# MoCaPS: A Machine Learning Model for Stratification of Cancer-Associated Cachexia Based on Blood Biomarkers

**DOI:** 10.64898/2025.12.23.25342866

**Authors:** Kayode D. Olumoyin, Magaret Park, Evan W. Davis, Jennifer B. Permuth, Katarzyna A. Rejniak

## Abstract

Identification of minimally invasive biomarkers of cancer-associated cachexia may help to recognize high risk patients for progression to more severe cachectic stages. We developed a machine learning-based <u>Mo</u>del for <u>Ca</u>chectic <u>P</u>atients <u>S</u>tratification (MoCaPS) to determine the sets of blood biomarkers that differentiate between noncachectic (NCa), precachectic (PCa), or cachectic (Ca) patients. The model was applied to data collected from treatment-naïve patients with pancreatic ductal adenocarcinoma through the Florida Pancreas Collaborative multi-institutional cohort study and biobanking initiative. Cachexia status of all participants was classified according to modified criteria by Vigano and colleagues. The MoCaPS model pipeline was designed to work effectively with datasets of moderate size to robustly select predictive data features, and to efficiently handle data imbalance. MoCaPS identified between 4 and 5 biomarkers out of 37 candidates that distinguished precachectic and cachectic stages, and demonstrated accuracies near or greater than 75% for predictors of NCa, PCa, and Ca.

## INTRODUCTION

Cancer-associated cachexia (CC) is a multifactorial syndrome observed in up to 80% of pancreatic ductal adenocarcinoma (PDAC) patients characterized by unintentional weight loss, muscle wasting in the presence or absence of fat loss, and fatigue [1], which can lead to a reduction in quality of life and poor clinical outcomes. [2, 3]. To distinguish between cachexia stages, we followed the Florida Pancreas Collaborative (FPC) and used criteria described in [4], which are based on the Vigano et al classification [5]. This classification system is comprised of the following types of data: (a) biochemistry (level of C-reactive protein (CRP) or albumin, or hemoglobin, or white blood cell count), (b) changes in food intake, (c) minimal or significant weight loss (WL), and (d) changes in daily activities based on the Patient-Generated Subjective Global Assessment (PG-SGA) performance status [6]. The recognized 4 cancer cachexia stages are: noncachexia (NCa), precachexia (PCa)—an early stage of the syndrome characterized by abnormal food intake or blood chemistry but no significant weight loss, cachexia (Ca), and refractory cachexia (RCa)—a stage that is largely irreversible [7]. However, several criteria used in the Vigano/FPC classification are based on patient-reported outcomes, which may be quite subjective, and thus differentiation between CC stages is difficult. This suggests the need for tools that can be based on more quantitative data, such as blood biomarker levels.

Moreover, there is also a dire need to develop minimally invasive approaches to identify CC earlier. Since blood is routinely collected clinically as a part of standard of care, identifying novel blood-based biomarkers of different stages of cachexia could be worthwhile. In previous work, certain blood biomarkers were deemed as prognostic for CC stages for PDAC patients [8-12]. These include CRP, interleukin-6 (IL-6), interleukin-8 (IL-8), tumor necrosis factor alpha (TNF-*α*), monocyte chemoattractant protein-1 (MCP-1), transforming growth factor beta (TGF-*β*), and growth/differentiation factor (GDF-15). Our previous work [12] also identified GDF-15 as a marker of CC that is predictive of survival, but only among Hispanic and Non-Hispanic White populations. However, these analyses use predominantly single feature correlation with the target outcome or pairwise data comparisons. Since CC is a complex multifactorial syndrome, there are potentially multidimensional and nonlinear interactions between different candidate biomarkers for CC. Single feature correlations may not fully capture these complex data relationships [13, 14]. In contrast, machine learning (ML) methods can successfully utilize multi-dimensional data focusing on patterns between numerous data features, and may identify data interconnections that yield non-intuitive predictions of target outcomes.

Developing tools that can predict early stages of cachexia may aid in earlier diagnosis of the disease and may allow for earlier therapeutic interventions. Such tools can also help clinicians in designing improved surveillance protocols for at-risk patients. In particular, differentiating between PCa and NCa stages is important for early detection of cancer patients who may not show symptoms of Ca (i.e. weight loss) but may be on a trajectory towards Ca status. This will benefit the patient, since interventions are more likely to be effective at the early stage.

We present here a novel ML-based framework (MoCaPS) that can handle nonlinear interconnectivities between patients’ blood-based biomarker data with the goal to identify a minimal set of biomarkers predictive of the different CC stages (NCa, PCa, or Ca). Our classifier has been applied to PDAC patients’ data collected by the FPC, a multi-institutional state-wide cohort study [15]. As a result, we propose three tools to distinguish between NCa vs. Ca stages, PCa vs. Ca stages, and PCa vs. NCa stages.

## RESULTS

### Computational study design

A total of 202 PDAC patients from the FPC [15] had available pre-treatment blood biomarker data [12] and CC status assessed using the modified Vigano et al. classification [4, 5]. The reported CC stages were along the cachexia continuum from NCa to PCa, to Ca, and to RCa. However, patients classified as RCa were excluded from this study due to small sample size and high pairwise positive correlations among several blood biomarkers (**Supplemental Figure S1**), which makes the RCa data insufficient for ML-based classification. After removing RCa cases, samples from 184 PDAC patients were used in our study, including 28 NCa, 53 PCa, and 103 Ca cases. For each patient, 37 blood biomarkers were considered (**Table 1**.)

**Table 1:**
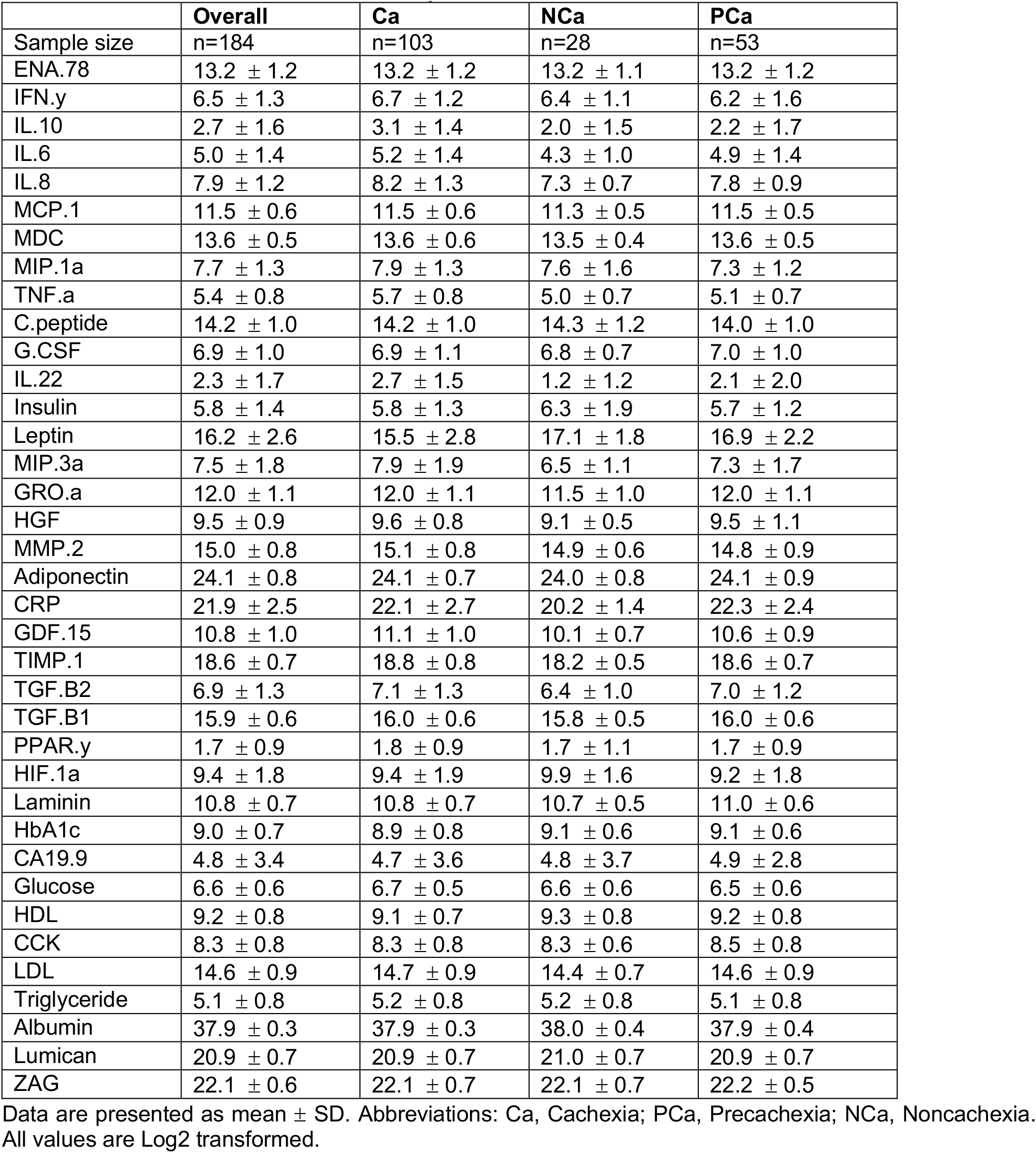
Blood biomarkers for PDAC patients from the Florida Pancreas Collaborative.

We developed three predictors that compared data for NCa vs. Ca or PCa vs. Ca, or PCa vs. NCa cachexia stages. Each predictor followed the MoCaPS pipeline shown in **Figure 1** that consists of four steps: (i) data preprocessing, (ii) feature selection, (iii) model training and testing, and (iv) generating model predictions and outcomes. The data preprocessing step includes splitting the given dataset of patients’ blood biomarkers into training and testing cohorts (70/30). Data in each cohort was then normalized using *MaxAbsScaler*, a class in the Scikit-learn Python library [16]. Next, the missing data were imputed using the multiple imputation with denoising autoencoders (MIDAS) method [17]. Finally, the correlated biomarkers were identified using the mutual information (MI) method [18] to generate an MI-based ranking of all features and pairwise correlation analysis to identify linear dependencies between features. The pairwise collinear features that had lower MI scores were removed. The feature selection step includes identification of a smaller subset of biomarkers that are robust in making prediction. This was achieved by applying the forward feature selection (FFS) method [19] with the feature importance score (FIS) [20, 21] to a normalized training dataset. Next, the sample size adequacy assessment was performed using a learning curve analysis [22, 23] to identify an appropriate ML classifier and to determine whether the training dataset is adequate for this classification task. We considered four classification models in this analysis: the support vector machines [24] with the radial basis function kernel (SVM-RBF), logistic regression (LR) [25], gradient boosting (GB) [26], and random forest (RF) [27] in order to determine the most suitable model for each considered classification. In the model training and testing step, the identified minimal set of biomarkers was used to learn hyperparameters for the chosen ML predictor. Subsequently, the optimal decision boundaries were determined using the Matthews correlation coefficient (MCC) method [28] to account for imbalanced datasets. In the model outcomes step, the performance of a given predictor was analyzed on the testing dataset and the confusion matrices and AUC/ROC curves were reported.

**Figure 1.**
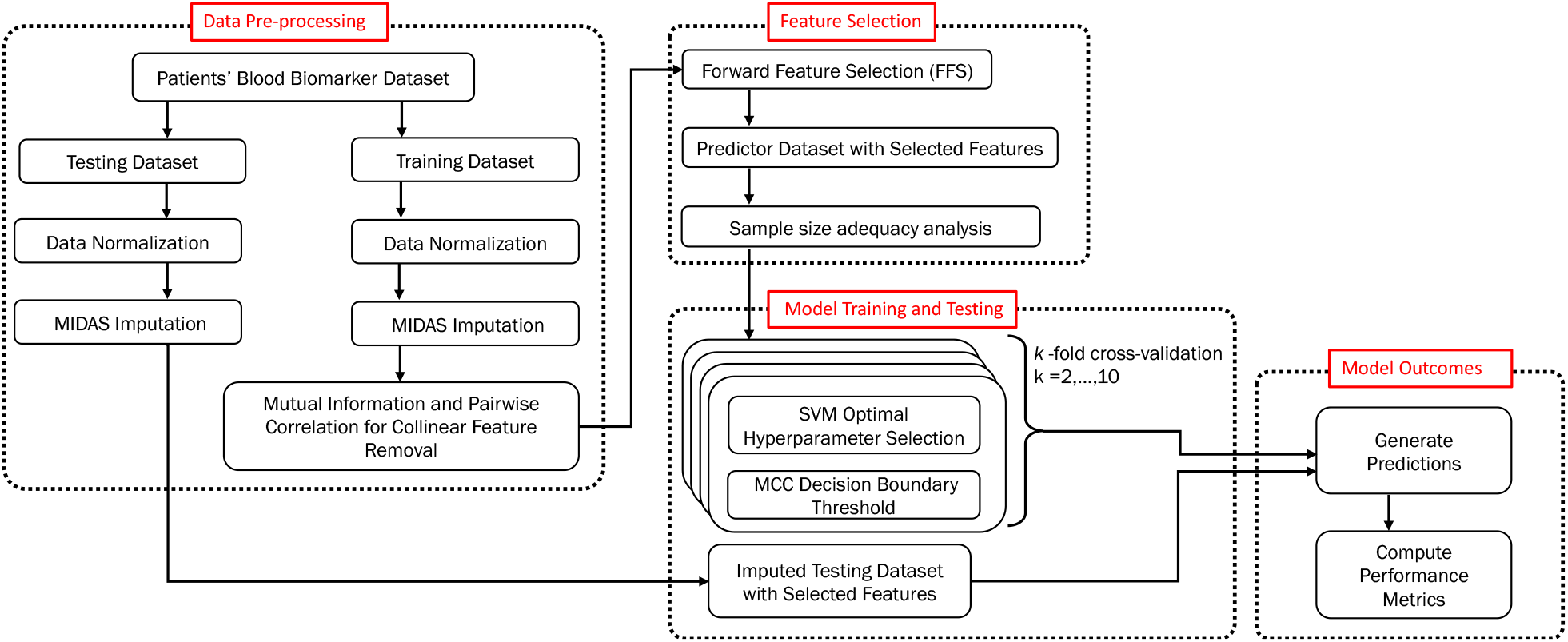
MoCaPS pipeline. Abbreviation: MoCaPS: Machine Learning **Mo**del for **Ca**chectic **P**atients **S**tratification based on blood biomarkers; MIDAS: Multiple Imputation with Denoising Autoencoders; FFS: Forward Feature Selection; SVM: Support Vector Machines; MCC: Matthews Correlation Coefficient.

The MoCaPS pipeline was implemented for three predictors: NCa vs. Ca, PCa vs. Ca, and PCa vs. NCa. In each case, MoCaPS identified a minimal list of predictive features, an ML classifier adequate to the given data, and the decision threshold for the imbalanced data. Finally, the performance metrics, including accuracy, sensitivity, and specificity, were computed for each of the three predictors.

### A computational predictor for stratification of non-cachectic vs. cachectic patients

131 PDAC patients’ data collected through the FPC biobank were classified either as Ca (103) or NCa (28). This dataset was split 70/30 into training (91 patients) and testing (40 patients) cohorts and each cohort was normalized using the *MaxAbsScaler* method. Each patients’ entry contained 31 blood biomarkers, however, for some cases the blood biomarkers values were missing, likely because they were outside the assessable range. The total proportion of missingness in this dataset was 1.5%, with 14 biomarkers having between 1 and 14 missing entries (**Supplemental Table S1**). The missing values were imputed using the MIDAS technique separately for the training and testing cohorts. The density plots of the training dataset and the training dataset with imputed values showed no significant differences between the data distributions in both datasets in each of the biomarkers with missing data (**Supplemental Figure S2**). The 37 biomarkers in the imputed training dataset were ranked according to their MI scores (**Figure 2A**) and the Pearson’s pairwise correlation coefficient was calculated for each pair of these biomarkers (**Figure 2B**). By setting the collinear threshold to 0.7 or higher (high positive correlation) and -0.7 or lower (high negative correlation), we identified pairs of biomarkers that met the collinearity threshold (**Figure 2C**). The following six biomarkers: ENA.78, IL.10, TNF.a, C.Peptide, IL.6, TIMP.1 (indicated in red in **Figure 2C**) met the collinearity threshold and were removed from further consideration due to lower MI scores. This reduced the dimension of the feature space from 37 to 31.

**Figure 2.**
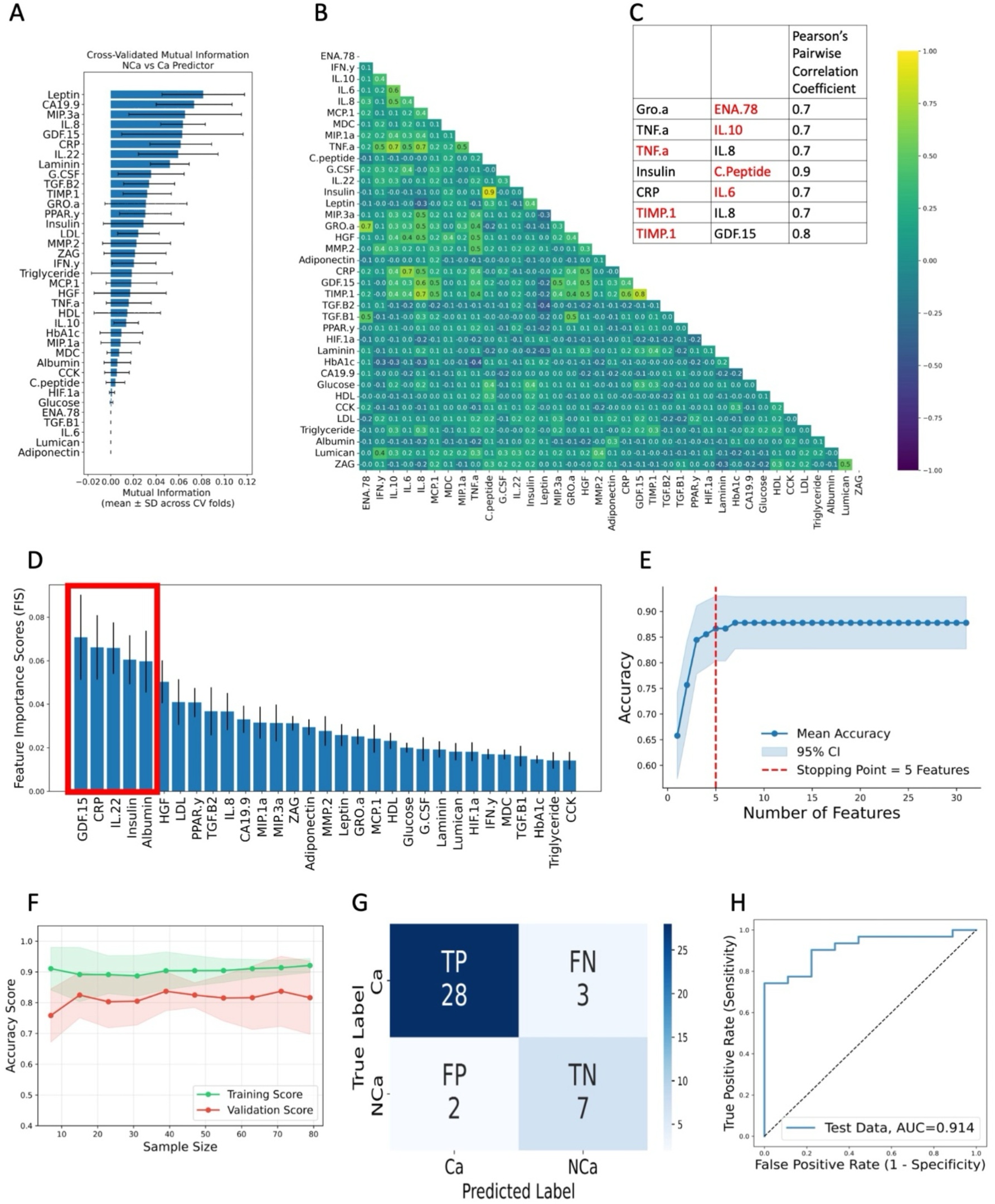
Identification of collinear biomarkers, feature selection and performance analysis for NCa vs. Ca predictor. **A.** MI score for all 37 biomarkers. **B**. Pearson’s pairwise correlation for all 37 biomarkers. **C**. A list of collinear biomarkers that met the cut-off threshold in **B** and a lower MI score in **A**; those indicated in red were removed from further analysis. **D**. The 31 candidate blood biomarkers pre-ranked using the FIS values. **E**. The cumulative curve of accuracy used to identify a subset of 5 robust predictive biomarkers indicated by the red box in **D. F**. The learning curves for training (green) and validation (red) across different proportions of the training dataset for the RBF-SVM predictor; the shaded regions represent standard deviation across 8-fold cross-validation. **G**. The confusion matrix of the RBF-SVM predictor and the corresponding performance metrics: Accuracy=0.875, Sensitivity=0.903, and Specificity=0.778. **H**. The AUC/ROC curve generated for predictions of NCa vs. Ca status for the testing set.

Using the imputed training cohort, a subset of the blood biomarkers that have high predictive accuracy in differentiating between NCa and Ca status was identified by applying the FIS ranking (**Figure 2D**). Next, the biomarkers were added sequentially in the order of pre-ranking to train the RF classifier, and its predictive accuracy was evaluated using the holdout validation set. This process yielded a monotonically increasing curve of the RF accuracy. The optimal number of biomarkers was determined using a stopping criterion that checks for a plateau in the validation accuracy curve [29], beyond which additional biomarkers provide minimal incremental gain in accuracy (**Figure 2E**). This method identified a set of 5 robust biomarkers (out of the initial set of 31) that together have a high predictive power. This set includes, according to their individual ranking: GDF-15, CRP, IL-22, Insulin, and Albumin (indicated by a red box in **Figure 2D**). The distributions of the predictive biomarker values in the training and testing cohorts are shown in **Supplemental Figure S3**.

Next, a sample size adequacy assessment was performed using the learning curve analysis [22, 23] to identify an appropriate ML classifier for the given task and the given data sample. The learning curves were obtained by stratified k-fold cross-validation on subsets of the training data containing from 10% to 100% of the available training dataset and evaluating performance on the held-out validation dataset using four different classifiers: RBF-SVM, LR, GB, and RF. We showed that for the 91-patient training dataset, RBF-SVM is a preferred classifier. The learning curves for the RBF-SVM presented in **Figure 2F** show high validation scores for most of the sampled subsets of the training dataset. The low variance between the training scores and the validation scores, as well as the near convergence in the validation scores indicate high generalization to new data and adequacy of the 91-training dataset for the classification task. The learning curves for the remaining three classifiers: LR, GB, and RF, are shown in **Supplemental Figure S4**.

For the identified classification method (RBF-SVM) and the 5 robust predictive biomarkers (GDF-15, CRP, IL-22, Insulin, and Albumin), the optimal hyperparameters (*C, γ*) were determined by using a *k*-fold cross-validation (for *k* = 2, …, 10) on the training cohort. Here, *C* specifies the width of the margins for avoiding data misclassification, and *γ* determines the nonlinearity of the decision boundary hyperplane. For each *k*, we used the MCC method to determine the best decision boundary threshold (*m*) that accounts for the imbalance in the training dataset. The optimal vales were *k*=6, *C* = 2.04, and *γ* = 0.060, and *m* = 0.60.

Finally, the model predictability was evaluated using the testing cohort with 40 patients’ data. The obtained confusion matrix of the model performance is shown in **Figure 2G**. The model generated an accuracy of 0.875, sensitivity (rate at which the model correctly predicts Ca) of 0.903, and specificity (the rate at which the model correctly predicts NCa) of 0.778. The area under the ROC curve (AUC) for the testing cohort was 0.914 (**Figure 2H**).

### A computational predictor for stratification of pre-cachectic vs. cachectic patients

In this case, there were 156 PDAC patients in the FPC database that were classified either as Ca (103) or PCa (53). This dataset was split 70/30 into training (109 patients) and testing (47 patients) cohorts. Each cohort was normalized using *MaxAbsScaler* method and missing entries were imputed using the MIDAS method. The observed proportion of missingness in the whole data set was 1.7%, with 16 biomarkers having between 1 and 18 missing entries (**Supplemental Table S2**). The density plots of the training dataset and the imputed training dataset have similar distributions for all biomarkers with missing data (**Supplemental Figure S5**). The 37 biomarkers were ranked according to their MI scores (**Figure 3A**) and Pearson’s pairwise correlation with the cut-off threshold of +/-0.7 were used to identify the collinear features (**Figure 3B**). Four biomarkers: Gro.a, TNF.a, IL.6, and TIMP.1 were removed from subsequent analysis because they met the collinearity threshold and had the lowest MI scores (**Figure 3C**). This effectively reduced the dimension of the feature space from 37 to 33.

**Figure 3:**
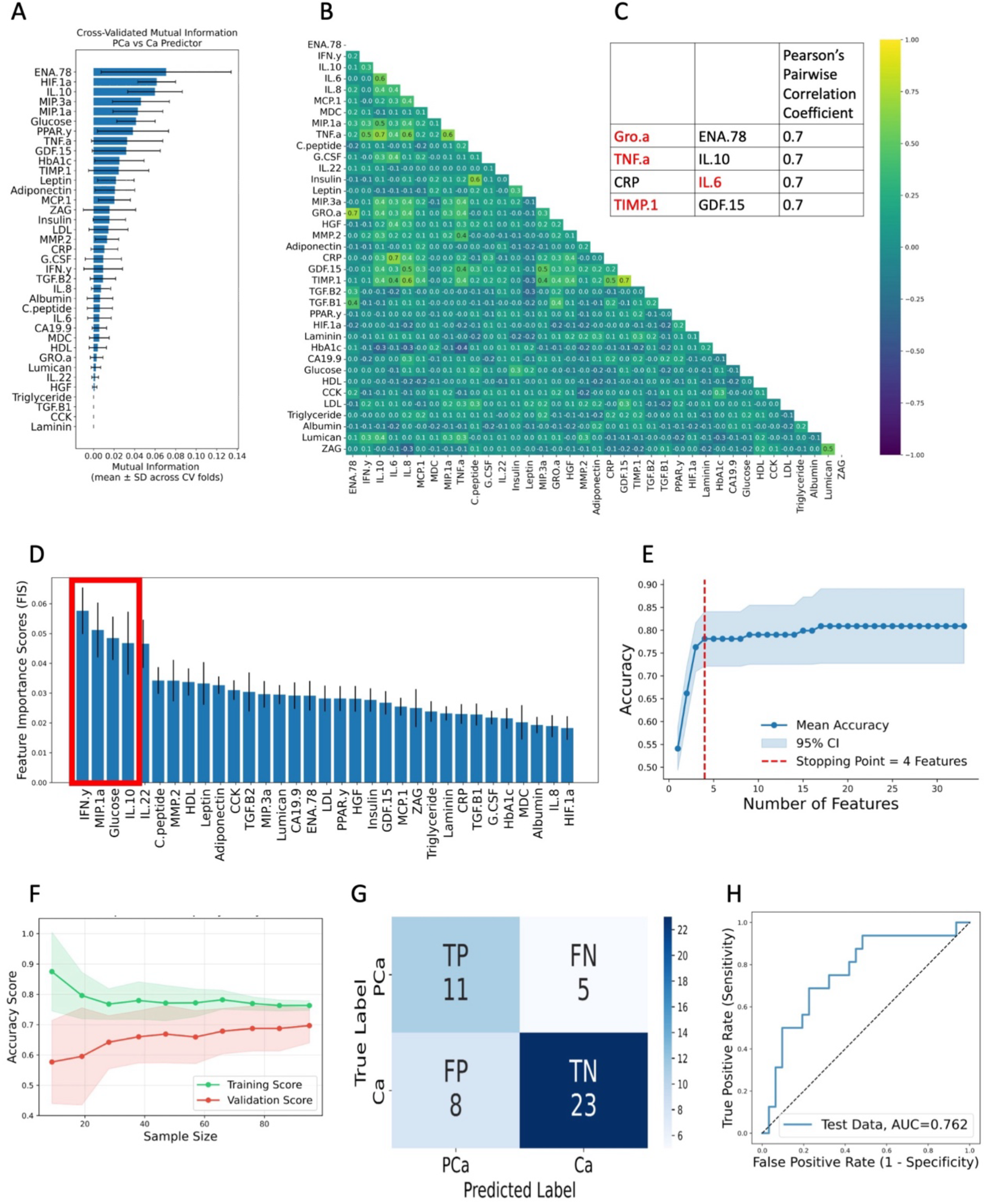
Identification of collinear biomarkers, feature selection and performance analysis for Ca vs. PCa predictor. **A.** MI score for 37 biomarkers. **B**. Pearson’s pairwise correlation for all 37 biomarkers. **C**. A list of collinear biomarkers from **B** with MI scores from **A;** those indicated in red were removed from further analysis. **D**. The 33 candidate blood biomarkers pre-ranked using the FIS values. **E**. The cumulative curve of accuracy used to identify a subset of 4 robust predictive biomarkers indicated by the red box in **D. F**. The learning curves for training (green) and validation (red) across different proportions of the training dataset for RBF-SVM predictor; the shaded regions represent standard deviation across 8-fold cross-validation. **G**. The confusion matrix of the RBF-SVM predictor and the corresponding performance metrics: Accuracy=0.723, Sensitivity=0.688, and Specificity=0.742. **H**. The AUC/ROC curve generated for predictions of Ca vs. PCa status for the testing set.

The training cohort with 33 blood biomarkers was used to identify a biomarker subset of high predictive accuracy in differentiating between Ca and PCa status. A ranking of the 33 blood biomarkers based on the FIS score was calculated using the RF method on a stratified cross-validated 10-fold subsampling of the training data (**Figure 3D**). Next, the contribution of individual biomarkers added sequentially in the order of their FIS ranking was assessed by obtaining a monotonically increasing curve of RF accuracy. Consequently, 4 out of the 33 biomarkers were identified as predictive (**Figure 3E**): IFN-*γ*, MIP-1*α*, Glucose, and IL-10 (**Figure 3D**). Distributions of the predictive biomarker values in the training and testing cohorts are shown in **Supplemental Figure S6**.

To analyze sample size adequacy and determine the best classification method, the learning curve analysis was employed on the 109-patient training dataset. Again, four classification methods were considered: RBF-SVM, LR, RF, and GB. Both RBF-SVM and LR outperformed the tree-based RF and GB classifiers (**Figure 3F** and **Supplemental Figure S7**) since they demonstrated low variance between the training scores and the validation scores indicating the adequacy of the given sample. In contrast, RF and GB overfit to the training data, likely due to the size of the training dataset. The RBF-SVM classifier was chosen over the LR method because of the absence of distinct thresholds in the Ca and PCa values in the selected biomarkers.

For the selected predictive biomarkers (IFN-*γ*, MIP-1*α*, Glucose, and IL-10) and the selected classification method (RBF-SVM), the optimal hyperparameters were identified (*C* = 1.100, *γ* = 1.240) together with the MCC optimal decision boundary threshold of *m* =0.283 using the training cohort. This classifier was evaluated on the testing cohort with 47 patients’ data, generating the confusion matrix shown in **Figure 3G**. The model yielded an accuracy of 0.723, sensitivity (rate at which the model correctly predicts PCa) of 0.688, and specificity (the rate at which the model correctly predicts Ca) of 0.742. The area under the ROC curve (AUC) for the testing cohort was 0.762 (**Figure 3H**).

### A predictor to differentiate between pre-cachectic and non-cachectic patients

For the PCa vs. NCa case, the FPC dataset contained 53 PCa patients and 28 NCa patients. This dataset was split 70/30 into training (56 patients) and testing (25 patients) cohorts. Data normalization, for each cohort separately, was performed using *MaxAbsScaler*. The total proportion of missingness in this dataset was (1.6%), with 12 biomarkers having between 1 and 11 missing entries (**Supplemental Table S3**). The density plots for comparison between the data distribution in the training dataset and the imputed training dataset show that they are indistinguishable in each of the biomarkers with missing data (**Supplemental Figure S8**). The MI ranking of all 37 biomarkers (**Figure 4A**) and Pearson’s pairwise correlation coefficient for those biomarkers (**Figure 4B**) identified two biomarkers: Insulin and TIMP.1 that met the collinearity threshold and had low MI scores (**Figure 4C**). These biomarkers were removed from further consideration, so that the feature space dimension was reduced from 37 to 35. The ranking of the 35 blood biomarkers based on FIS scores computed through the RF method on a stratified cross-validated 10-fold subsampling of the training data is shown in **Figure 4D**. Subsequently, the FFS method yielded a monotonically increasing curve of RF accuracy showing the contribution of sequentially added biomarkers in the order of their FIS ranking. This identified 4 out of the 35 biomarkers to be predictive in differentiating between NCa and PCa status (**Figure 4E**). The selected biomarkers were: CRP, MIP-3*α*, GRO.*α*, and ZAG. Their distributions in the training and testing cohorts are presented in **Supplementary Figure S9**.

**Figure 4:**
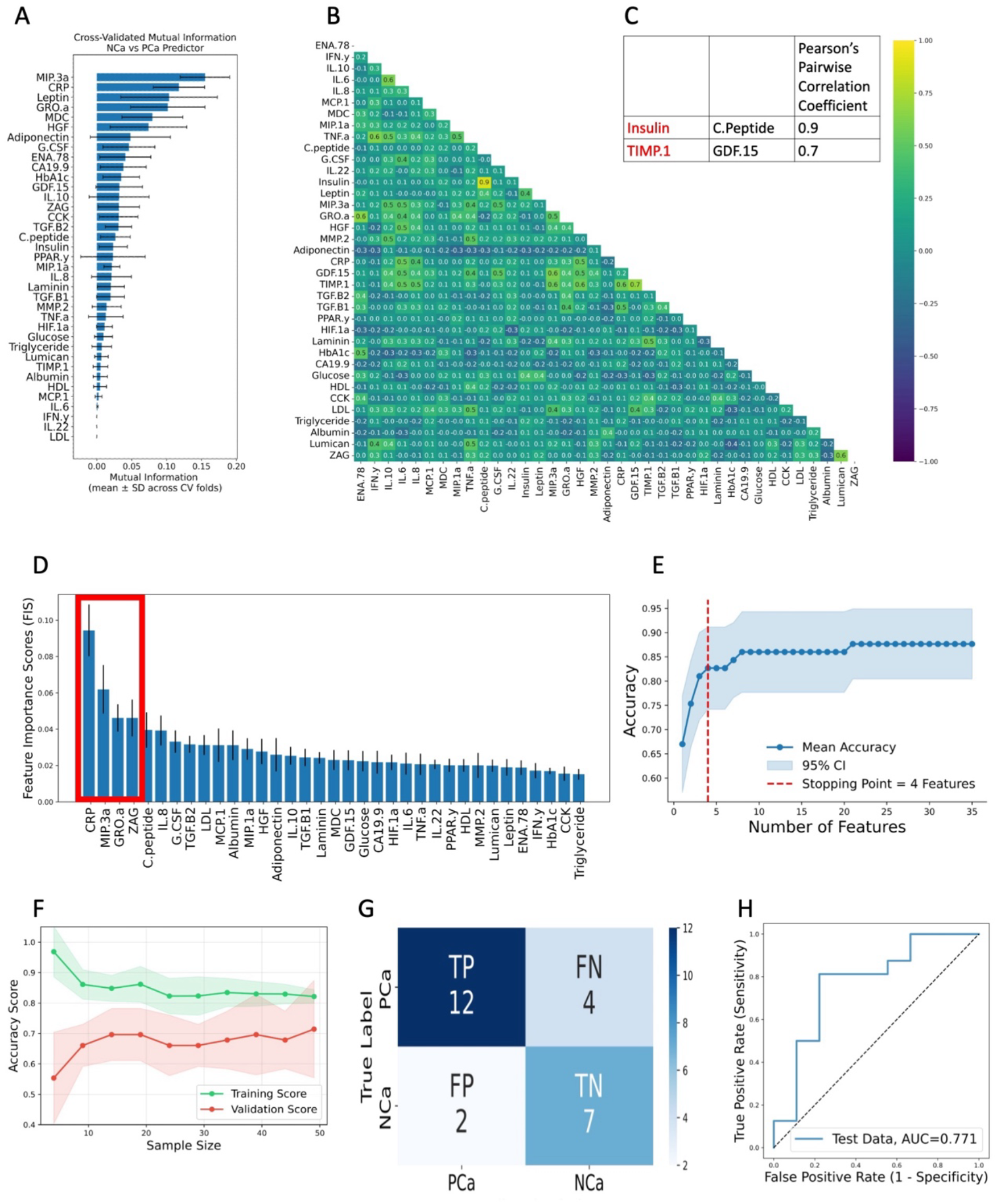
Identification of collinear biomarkers, feature selection and performance analysis for NCa vs. PCa predictor. **A.** Mutual Information score for all 37 biomarkers. **B**. Pearson’s Pairwise correlation for all 37 biomarkers. **C**. The collinear biomarkers from **B** that met the cut-off threshold and had a lower MI score in **A** were removed from further consideration; these biomarkers: ‘Insulin’, ‘TIMP.1’ are indicated in red. **D**. The 35 candidate blood biomarkers were pre-ranked using the FIS values. **E**. The cumulative curve of accuracy identified a subset of 4 robust predictive biomarkers indicated by the red box in **D. F**. Learning curves for training (green) and validation (red) across different proportions of the training dataset. Shaded regions represent standard deviation across 8-fold cross-validation. **G**. The confusion matrix of the RBF-SVM predictor and the corresponding performance metrics: Accuracy=0.760, Sensitivity=0.750, and Specificity=0.778. **H**. The AUC/ROC curve generated for predictions of NCa vs. PCa status for the testing cohort.

To assess size adequacy of the 56-patient training dataset and identify the best classification method, we used the learning curve analysis method for four classification methods: RBF-SVM, LR, RF, and GB. Both RBF-SVM and LR showed reasonably good performance on the training dataset (**Figure 4F** and **Supplemental Figure S10**) and were prioritized over the tree-based classifiers: RF and GB. RF and GB demonstrated high variance between validation scores and training scores, a high indication of overfitting to the training data (**Supplemental Figure S10**). RBF-SVM and LR, on the other hand, both demonstrated low variance between the training scores and the validation scores. The convergence trend of the validation scores in RBF-SVM and LR showed that model performance can probably be improved with additional data. The lack of distinct thresholds in the NCa and PCa values in the selected biomarkers (**Supplemental Figure S9**) also means that RBF-SVM was chosen over LR for final stratifications.

For the four predictive biomarkers (CRP, MIP-3*α*, GRO.*α*, and ZAG) and the selected classification method (RBF-SVM), the optimal hyperparameters (*C* = 1.350, *γ* = 1.130) were identified together with the optimal decision boundary threshold of *m* =0.647 using the training cohort. This classifier was evaluated using the testing cohort of 25 patients and the obtained confusion matrix is shown in **Figure 4G**. The model yielded an accuracy of 0.760, sensitivity (rate at which the model correctly predicts PCa) of 0.750, and specificity (the rate at which the model correctly predicts NCa) of 0.778. The area under the ROC curve (AUC) for the testing cohort was 0.771 (**Figure 4H**).

## DISCUSSION

In this study, we aimed to use blood biomarker data to develop machine-learning predictors in order to differentiate between patients’ cachexia stages. We utilized data collected by the Florida Pancreas Collaborative and the CC classification criteria that were modified from the Vigano et al. system [4, 5, 12]. Because there are potentially complex interactions between different candidate blood biomarkers for CC, we focused on identifying a minimal set of biomarkers that together had predictive power. For each of the three classification tasks (NCa vs. Ca, Ca vs. PCa, and NCa vs. PCa), the forward feature selection method narrowed the number of predictive biomarkers to 4-5 that were optimal in distinguishing between two different cachexia stages. We also determined that the training dataset sample size was adequate to use the RBF-SVM classifier (with MCC adjustment) in each of the three considered cases. In particular, a set of 5 biomarkers: GDF-15, CRP, IL-22, Insulin, and Albumin was optimal in distinguishing between NCa vs. Ca stages with the accuracy of 87.5%. In the case of Ca vs. PCa, we found the following set of 4 biomarkers to be predictive when used together: IFN-*γ*, MIP-1*α*, glucose, and IL-10, yielding the accuracy of 72.3%. Moreover, we demonstrated that a set of 4 biomarkers: CRP, MIP-3*α*, GRO.*α*, and ZAG can together distinguish the NCa status from PCa with the accuracy of 76%. In all three cases, the classifiers specific was between 74% and 77.8%, and the AUC values were between 76.2% and 91.4%. Among the identified predictive blood biomarkers used in all three predictors, only CRP overlapped between the predictive biomarkers in the NCa vs. Ca and NCa vs. PCa predictors.

We recognize that CRP is also used as one of the criterium of the modified Vigano system for CC classification [4, 5, 12]. However, there is no one-to-one correlation between CRP levels and CC status, since this criterium is complex and contains thresholds for levels of either CRP or albumin, or hemoglobin, or white blood cell count. Similarly, in our classification, CRP is one of the biomarkers with multidimensional and nonlinear interactions that have predictive value only in combination with 3 or 4 other blood biomarkers. Moreover, several blood-based biomarkers that were previously reported to correlate with CC status for pancreatic cancer patients in our studies and by others [8-12], were also identified as predictive in our approach. These include CRP and GDF-15, however another two—interleukin-6 (IL-6) and interleukin-8 (IL-8)—were not selected as necessary in any of our three predictors.

In our previous work [12], we analyzed the AUC for several analytes which were significantly different between NCa and Ca patient groups. These blood-based biomarkers included WBC count, albumin, and hemoglobin that are typically associated with cachexia status, as well as GDF-15 and TNF-α that were identified as significantly higher in patients with Ca compared with those with NCa. For WBC count, albumin, and hemoglobin, the AUC values were between 57% and 63%, for GDF-15 or TNF-α alone, or both combined, the AUC values were between 71% and 76% [12]. However, our ML predictor that uses a combination of multiple blood-based biomarkers shows AUC of 91.4% which indicates that by considering more complex interconnectivity between patients’ blood-based biomarkers may increase their predictability. There are a few published studies that used machine learning-based approaches to identify possible biomarkers for Ca from clinical data. In [30], the authors used demographic, clinical, and patient reported outcomes (PRO) from a multi-center patient cohort study to identify biomarkers that predict Ca and PCa status. One of the factors identified to be predictive was the C-reactive protein (CRP), which was also selected by our approach. The model developed in [30] reported an AUC value similar to our study for differentiation between NCa vs. Ca (∼0.83). Similarly, *CRP* was identified as one of the 15 top predictive biomarkers of Ca in the case when weight loss information is not available [31]. The data used by this ML-based model consisted of demographic, cancer-related clinical data, PRO related to GI symptoms, and blood-based biomarkers. The model showed good performance for predicting Ca in the validation set with the AUC of 0.763. However, this model did not address PCa status. Another ML-based approach by the same authors was used to predict potentially reversible cancer cachexia, which was defined as a cachexia diagnosis at baseline that turned negative one month later [32]. This model used clinical and demographic data for 16 different tumors, but no blood-based biomarkers. This model showed very good predictability with the AUC of 0.887 for the holdout test set and AUC of 0.863 for the external validation set. It was also suggested that the generated results can provide insights into symptoms that can be addressed to prevent or treat PCa.

One of the limitations of this study is lack of independent dataset for external validation. Since several of blood-based biomarkers used in our predictors are not collected as a part of the standard of care procedures for the PDAC patients, further studies are needed to identify a suitable dataset to validate our findings externally. Moreover, the machine learning classifier presented here was used to stratify patients’ blood biomarkers data into different stages of cachexia based on the modified Vigano system [4, 12]. However, similar computational frameworks can be developed for other cachexia classification criteria, such as Fearon et al. [33], Vigano et al. [5], or Martin et al. [34].

In summary, this study showed that the integration of machine learning and deep learning techniques, such as multiple imputation method to handle missing data, feature selection techniques to identify a minimal predictive subset of blood biomarkers, and machine learning classification that incorporates algorithms for handling data imbalance and cross-validation for optimal hyperparameter tuning, can identify predictive blood-borne biomarkers and stratify PDAC patients into different cachexia stages. Thus, the developed method has a high translational potential and could be used as a supportive tool in earlier diagnosis of the disease for cancer patients who may not show symptoms but may be on a trajectory towards CC. It may also aid in improving supportive care and clinical outcomes in the treatment of PDAC patients who are at risk of CC. Finally, it can be used as part of surveillance strategy for patients at risk of progressing to a more severe cachectic stage.

Our study provided three novel ML models to differentiate between cachexia stages based on blood biomarkers, that are minimally invasive and easily accessible. These predictors work well with the moderate datasets and yield favorable performance metrics. These predictors may be incorporated into clinicians’ diagnostic tools for detecting early-stage cachexia and assessing the risk of progressing to a more severe cachectic stage.

## METHODS

### Study population and data collection

This study included patient data collected by the Florida Pancreas Collaborative (FPC), a multi-institutional prospective cohort study and biobanking initiative between 2018 and 2021, and approved by the Moffitt Cancer Center Scientific Review Committee (MCC19717, Pro00029598), and Advarra IRB (IRB00000971). All patients provided informed consent for participation [15]. Pre-treatment serum biomarker levels that comprised of cytokines, chemokines, adipokines, lipoproteins, glycans, and other analytes (37 biomarkers in total) were available for 202 patients [12]. Patients’ CC stage was determined using the modified Vigano criteria [4, 5] and categorized into 4 CC stages: NCa, PCa, Ca, and RCa. However, due to low number of cases (n=18) unsuitable for ML algorithms, patients with RCa status were excluded from this study.

### Description of the ML classification problem

Our goal was to identify a predictive subset of features that differentiates between two targets in a binary classification task, and to provide metrics of success for such data stratification. Let the dataset ***X*** consists of *M* data points and *P* features: ***X*** = [***X***_1_, ***X***_2_, ⋯, ***X***_*M*_]^⊺^, where each data point is 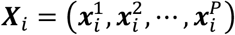 for *i* ∈ {1,2, ⋯, *M*}. Additionally, for each *i*, the corresponding target is the binary class *y*_*i*_ ∈ {−1, 1}. We considered three different machine learning predictors: NCa vs. Ca, PCa vs. Ca, and PCa vs. NCa. In each case, the goal was to find the minimal subset of features of size *Q*, where *Q* < *P*, that divides the dataset ***X*** into distinct binary classes. We used a data-informed approach: First, we split ***X*** into training and testing cohorts; both cohorts were normalized and the missing data were imputed. Using the training cohort, we removed the correlated features and implemented feature selection method to identify predictive features. Next, we assessed sample size adequacy and determined an optimal machine learning classifier. For that classifier, the optimal hyperparameters were found by using a cross-validation technique and the optimal decision boundary threshold was learnt to correct for imbalance in ***X***. Finally, this classifier was applied to the testing cohort to assess the prediction metrics.

### Data normalization

Data normalization was performed to ensure that all features can contribute equally. In order to prevent data leakage between training and testing cohorts, we split the overall data before data normalization, and performed normalization on the training and testing cohorts separately. We used the *MaxAbsScaler*, a class in the scikit-learn Python library [16] which translates each feature independently to have a maximal absolute value of 1.0, preserving data distribution but only linearly scaling down each biomarker.

### Multiple imputation framework for data retention

Multiple imputation is a statistical method to handle missing data. Let ***X*** ∈ ℝ^*M*×*P*^ be a data matrix with observed entries ***X***_*obs*_ and missing entries ***X***_*miss*_. Under the assumption that data are missing at random (MAR) or completely at random (MCAR), the multiple imputation replaces all entries in ***X***_*miss*_ with imputed values that preserve the interrelations in ***X***_*obs*_. We used the multiple imputation with denoising autoencoders (MIDAS) method [17], which is a scalable deep learning-based technique that employs a class of unsupervised neural networks known as denoising autoencoders [35] and Monte Carlo dropout to generate multiple imputation of the missing data with realistic uncertainty quantification. MIDAS was used separately for each of the three predictors (NCa vs. Ca, Ca vs. PCa, and NCa vs. PCa), and in all cases the distributions for each biomarker for the training and the imputed training datasets were compared.

### Mutual information measure for nonlinear dependencies

Mutual information (MI) is a non-parametric measure of statistical dependency between the dataset ***X*** ∈ ℝ^*M*×*P*^ and the predicted binary class ***Y*** ∈ ℝ^*M*×1^, where *M* is the number of patients and *P* is the number of features. MI captures nonlinear dependencies in high-dimensional data, which makes them robust for measuring feature relevance in discrete or categorical datasets [36, 37]. A discrete MI is computed as follows: 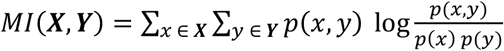. Where *p*(*x, y*) is the joint probability of ***X*** and ***Y***, while *p*(*x*) and *p*(*y*) are the marginal probabilities of ***X*** and ***Y***, respectively.

### Forward feature selection method for identification of predictive blood biomarkers

The feature selection was performed to identify the minimal predictive subset of features for each data classifier, and to reduce the number of biomarkers used in each case. Using the Forward Feature Selection (FFS) method [19], we built the stratified cross-validated 10 subsamplings of the training dataset, taking 70% of the data at a time and partition them into training and holdout sets. On each of the 10 subsamples, we trained a random forest (RF) classifier and obtained the feature importance score (FIS) [20, 21] ranking for each of the biomarkers. Next, the biomarkers were added sequentially in the order of pre-ranking to train the RF classifier, and its predictive accuracy was evaluated using the holdout validation set. This process of adding biomarkers one at a time yields a non-strictly increasing curve of the RF accuracy. There may be local regions of downward fluctuations due to noisy biomarkers. To mitigate these fluctuations, accuracy curves were converted to a monotonically increasing curve, retaining the maximum accuracy observed up to each biomarker addition step. The optimal number of features for each fold was determined using an elbow-point detection method [29, 38, 39], which identifies the point beyond which additional biomarkers provide minimal incremental gain in accuracy. This stopping criterion also prevents overfitting to the training data. The mean accuracy curve and 95% confidence interval across all folds were used to visualize biomarker contribution to the predictiveness of the model and to determine final biomarker selection. This approach eliminates multicollinearity of blood biomarkers for prediction purposes.

### Learning curve for determining sample size adequacy

To assess training dataset adequacy for a classification task of distinguishing between cachectic stages, we performed learning curve analysis using stratified k-fold cross-validation, where we repeatedly subsampled the training data at different sample sizes. We trained four models: Logistics Regression [25], Random Forest [27], Gradient Boosting [26], and Radial Basis Function-Support Vector Machine [24] on subsets of the training dataset from 10% to 100% of the available training dataset and evaluated performance on the holdout validation dataset.

### Support vector machine model for learning the optimal classification rules

For each data point 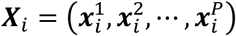 in ***X*** ∈ ℝ^*M*×*P*^, a binary ML classifier was used to learn a corresponding prediction *y*_*i*_ in ***Y*** ∈ ℝ^*M*×1^. The accuracy of this prediction depends on how well the classifier identifies an optimal decision boundary that separates ***X*** into the distinct binary classes. However, the interactions between the predictive features are often multidimensional and nonlinear. A support vector machine (SVM) model with the nonlinear Radial Basis Function (RBF) kernel: (.***X***_*i*_, ***X***_*j*_) = *exp* (−*γ* ‖ ***X***_*i*_ − ***X***_*j*_ ‖^2^) [40, 41] can potentially capture these complex interactions between features as it learns to distinguish between the distinct classes in the training dataset and generalize to a new dataset. First, all data was split 70/30 into the training and testing cohorts. Then, the optimal RBF-SVM hyperparameters (*C, γ*) were identified by performing *k*-fold cross-validation (CV) on the training dataset, for *k* = 2, …,10. Here *C* specifies the width of the margins for avoiding data misclassification and *γ* controls the nonlinearity of the decision boundary hyperplane. For each *k*, the best *C* and *γ* were determined by (i) minimizing the difference between the cross-validation training accuracy and testing accuracy; (ii) maximizing the cross-validation training accuracy; and (iii) minimizing the value of *C*. The optimal values of *C* and *γ* were obtained by computing the Matthews correlation coefficient (MCC) for each cross-validation (*k* = 2, …,10) and then choosing *C* and *γ* for which MCC is maximal, and *C* is minimal.

### Matthews correlation coefficient algorithm to account for data imbalance

The Matthews correlation coefficient (MCC) statistical test [28, 42] was used to determine the optimal decision boundary threshold to account for true and false positives and negatives in the imbalanced training dataset. Usually, this threshold is set to 0.5, because it is assumed that SVM works with balanced data (i.e., similar numbers of data in each class). For the imbalanced dataset, this threshold has to be adjusted. For learnable parameters ***w***, *b*, the magnitude of the decision function ***w***^⊺^***X***_*i*_ + *b* for each ***X***_*i*_ was extended to probability using the Scikit-Learn library [16]. Next, the thresholds between 0.1 and 0.9 were tested by separating the training cohort data into binary classes (*Positive* or *Negative*) based on whether their RBF-SVM–generated prediction probabilities exceeded the given threshold. For each threshold, MCC was calculated on the labeled prediction probabilities. The threshold with the maximum MCC was used as the decision boundary threshold to generate predictions on the testing cohort. A detailed algorithm is presented in **Supplemental Algorithm S1**.

### Performance metrics

To assess performance of the classification protocol on the testing dataset, the following performance metrics were used for evaluation: (1) true positive rate (TPR) or sensitivity, is the percentage of correctly classified positive instances: TPR=TP/(TP+FN); (2) true negative rate (TNR) or specificity, is the percentage of correctly classified negative instances: TNR=TN/(FP+TN); (3) accuracy is the percentage of correctly classified positive and negative instances: accuracy=(TP+TN)/(TP+FN+FP+FN); (3) area under the receiver operating characteristics curve (AUC/ROC or AUC) measures the ability to discriminate between positive and negative cases and ranges from 0.5 (coin toss) to 1.0 (perfect classification), when ROC curve shows tradeoffs between TP and FP. Here, TP (true positive) is the correctly classified data, TN (true negative) is the correctly classified data, FP (false positive FP) is the misclassification of the positive class, and FN (false negative) is the misclassification of the negative class.

## Data and code availability statement

The data and code used in this study is available from the following depositories: github.com/okayode/MoCaPS and github.com/rejniaklab/MoCaPS

## Data Availability

The data and code used in this study is available from the following depositories:
github.com/okayode/MoCaPS and github.com/rejniaklab/MoCaPS

https://github.com/okayode/MoCaPS

https://github.com/rejniaklab/MoCaPS

## ACKNOWLEDGMENT

This work was supported in part by the Department of Defense Health Program Congressionally Directed Medical Research Program grants: W81XWH-22-1-0340 (to KAR), and W81XWH-22-1-1021 LOG#PA210192 (to JBP), by the US National Institutes of Health, National Cancer Institute grant R01-CA259387 (to KAR), and by the James and Esther King Biomedical Research Program, Florida Department of Health grants #8JK02 and #24K03 (to JBP). This work was supported in part by the Shared Resources at the H. Lee Moffitt Cancer Center & Research Institute an NCI Designated Comprehensive Cancer Center under the grant P30-CA076292 from the National Institutes of Health. The funders played no role in study design, data collection, analysis, and interpretation of data. All data used in this computational study was provided by the Florida Pancreas Collaborative (FPC), a state-wide initiative and biobank. We would like to thank all participating FPC institutions, members, and patients.

## AUTHORS CONTRIBUTIONS

KDO and KAR conceptualized the project. KDO designed the presented work, developed the computational model, and created the software. JBP, EWD, and MP prepared and provisioned the data and provided recommendations on interpretation of the results. KDO and KAR drafted the manuscript. All authors revised the manuscript.

## ETHICS APPROVAL

This study was conducted according to guidelines of the Declaration of Helsinki, and approved by the Moffitt Cancer Center Scientific Review Committee (MCC19717, Pro00029598), and the Institutional Review board Advarra IRB (IRB00000971). All patients provided informed consent for participation.

