## Supplementary Material for "MoCaPS: A Machine Learning Model for Stratification of Cancer-Associated Cachexia Based on Blood Biomarkers"

#### Supplementary Information

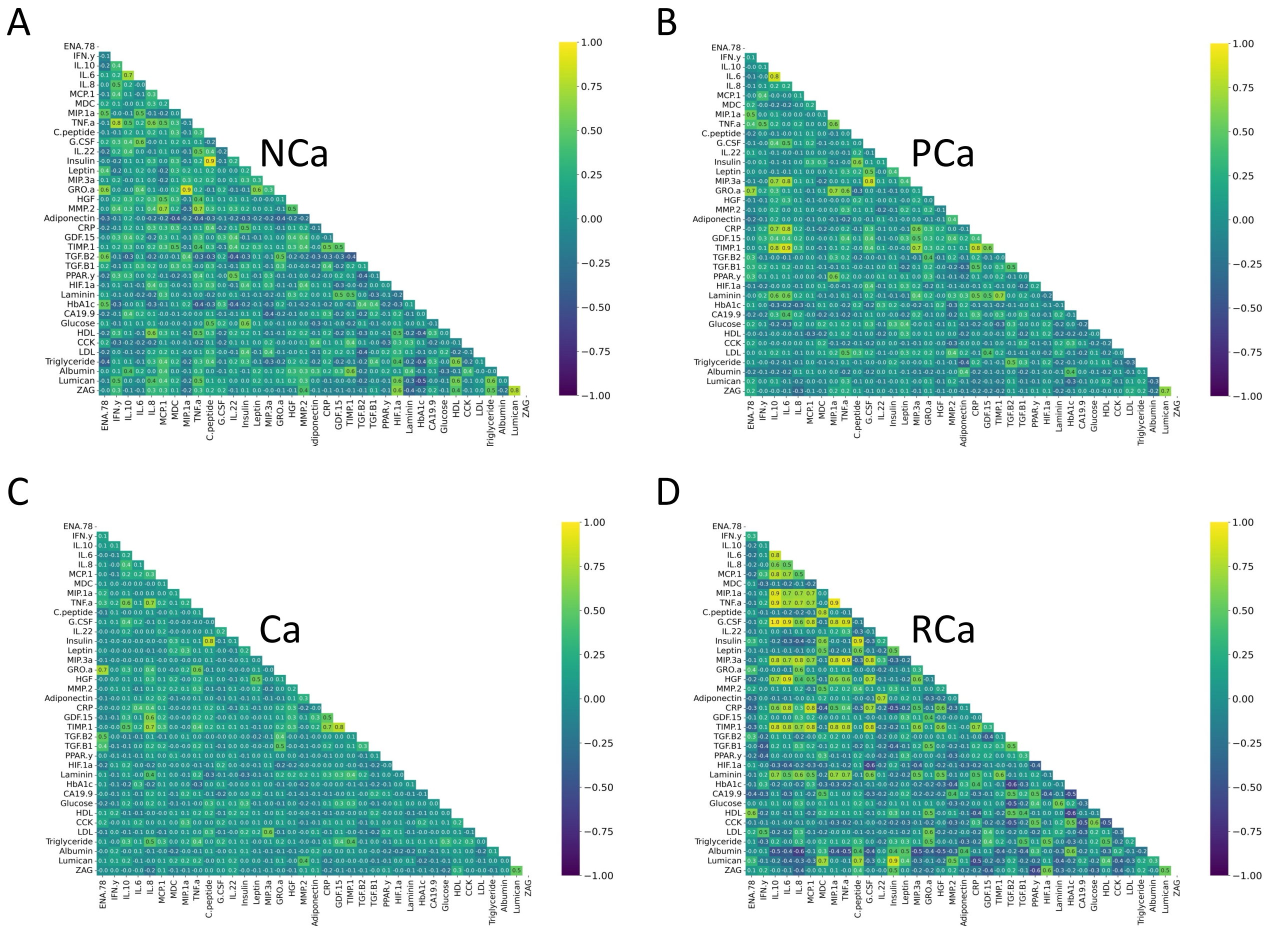


**S1 Figure**: Pairwise correlation heatmaps between blood biomarkers from the entire dataset for: **A**. noncachexia (NCa) dataset, **B**. precachexia (PCa) dataset, **C**. cachexia (Ca) dataset, **D**. refractory cachexia (RCa) dataset. In the RCa case, a sizeable number of the biomarkers was positively correlated (correlation coefficient greater than 0.70), which makes the RCa data unreliable for a machine learning classification task. In the other 3 cases, the biomarkers were not highly correlated.

**S1 Table:** The number of patients with missing data for the NCa vs. Ca cohort of 131-patients and 37 biomarkers

| | Missing |
|---|---|
| ENA.78 | 0 |
| IFN.y | 0 |
| IL.10 | 0 |
| IL.6 | 0 |
| IL.8 | 0 |
| MCP.1 | 0 |
| MDC | 0 |
| MIP.1a | 4 |
| TNF.a | 0 |
| C.peptide | 0 |
| G.CSF | 0 |
| IL.22 | 5 |
| Insulin | 1 |
| Leptin | 3 |
| MIP.3a | 8 |
| GRO.a | 0 |
| HGF | 0 |
| MMP.2 | 2 |
| Adiponectin | 0 |
| CRP | 0 |
| GDF.15 | 0 |
| TIMP.1 | 0 |
| TGF.B2 | 14 |
| TGF.B1 | 0 |
| PPAR.y | 1 |
| HIF.1a | 10 |
| Laminin | 0 |
| HbA1c | 0 |
| CA19.9 | 9 |
| Glucose | 4 |
| HDL | 0 |
| CCK | 1 |
| LDL | 0 |
| Triglyceride | 13 |
| Albumin | 1 |
| Lumican | 0 |
| ZAG | 0 |

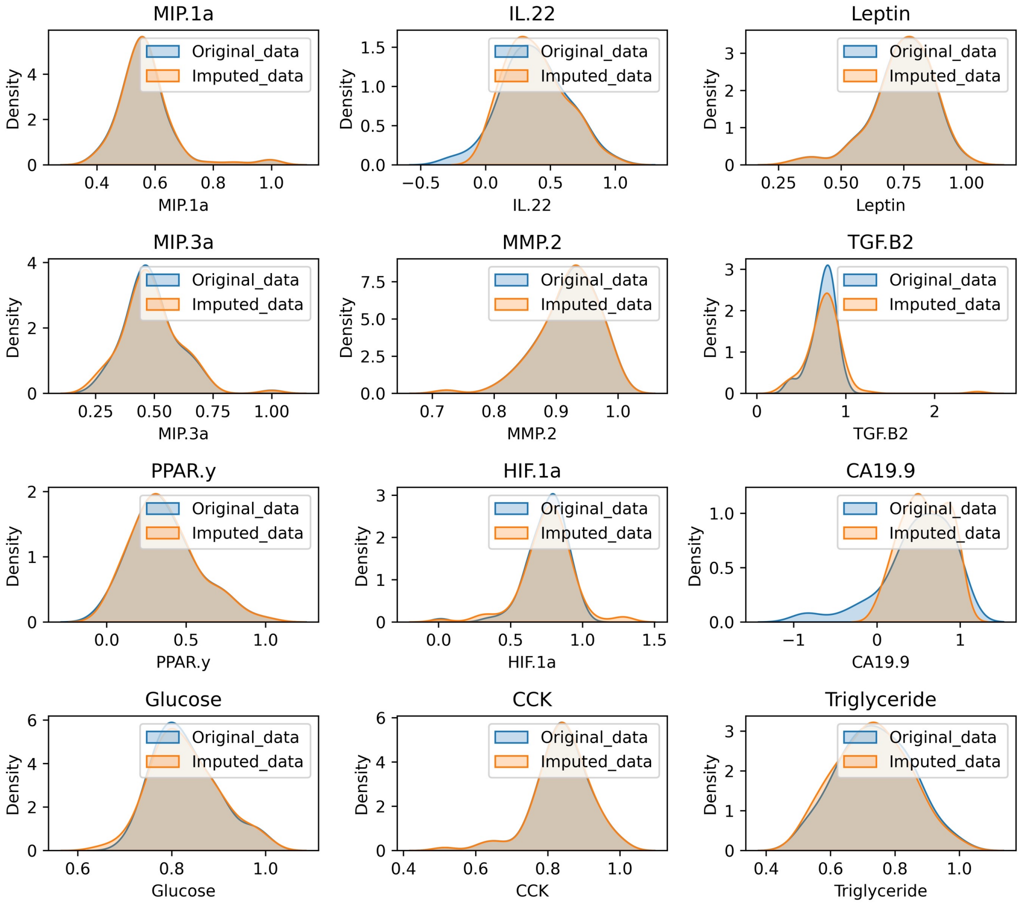


**S2 Figure:** Density plots of the training dataset (blue) and the imputed training dataset (orange) for 12 biomarkers with missing data for the NCa vs. Ca predictor.

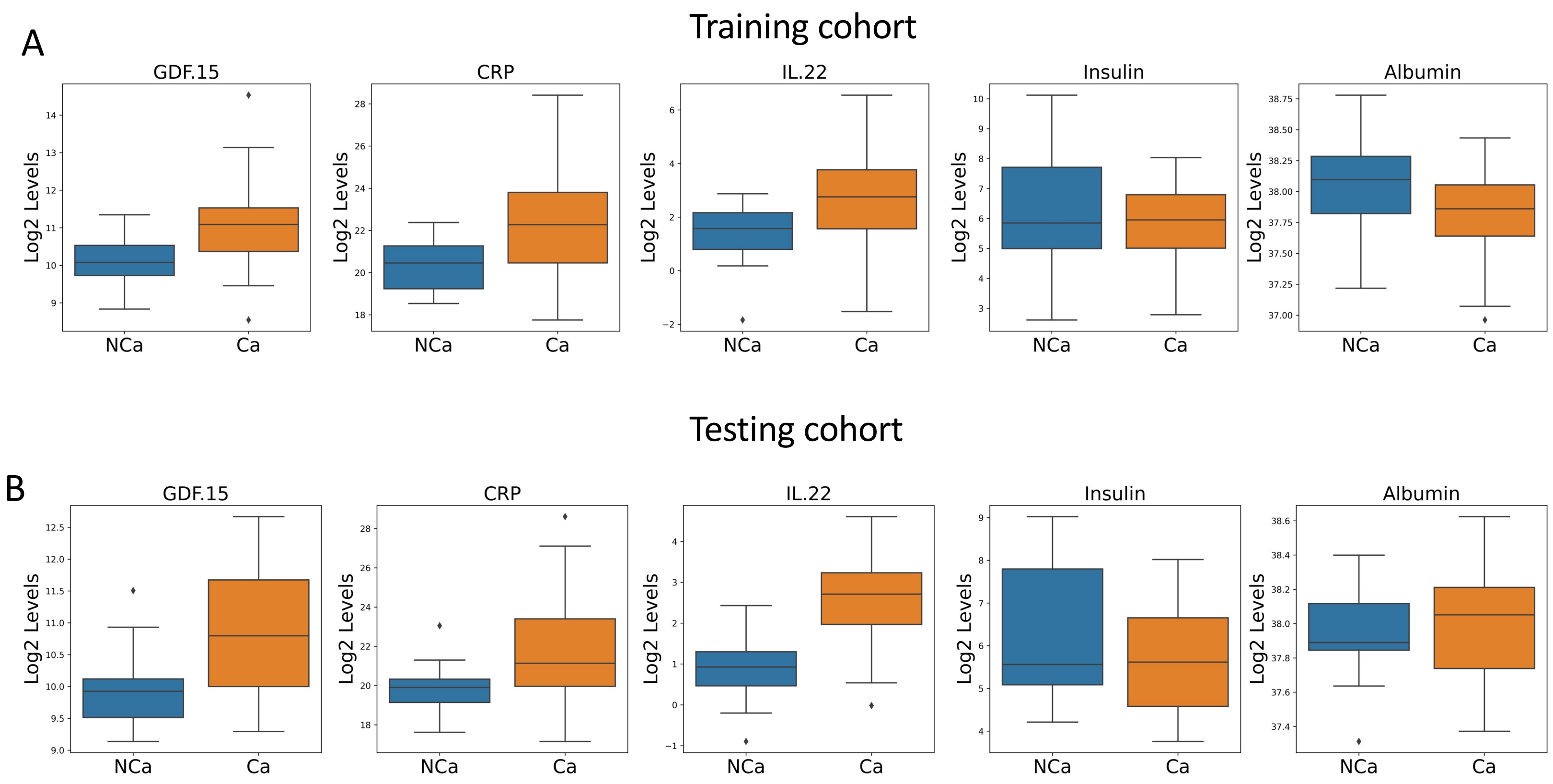


**S3 Figure: Distributions of robust predictive biomarkers for the NCa vs. Ca predictor.** Distributions of the NCa (blue) and Ca (orange) classes for the training **A** and testing **B** cohorts for the five robust predictive biomarkers.

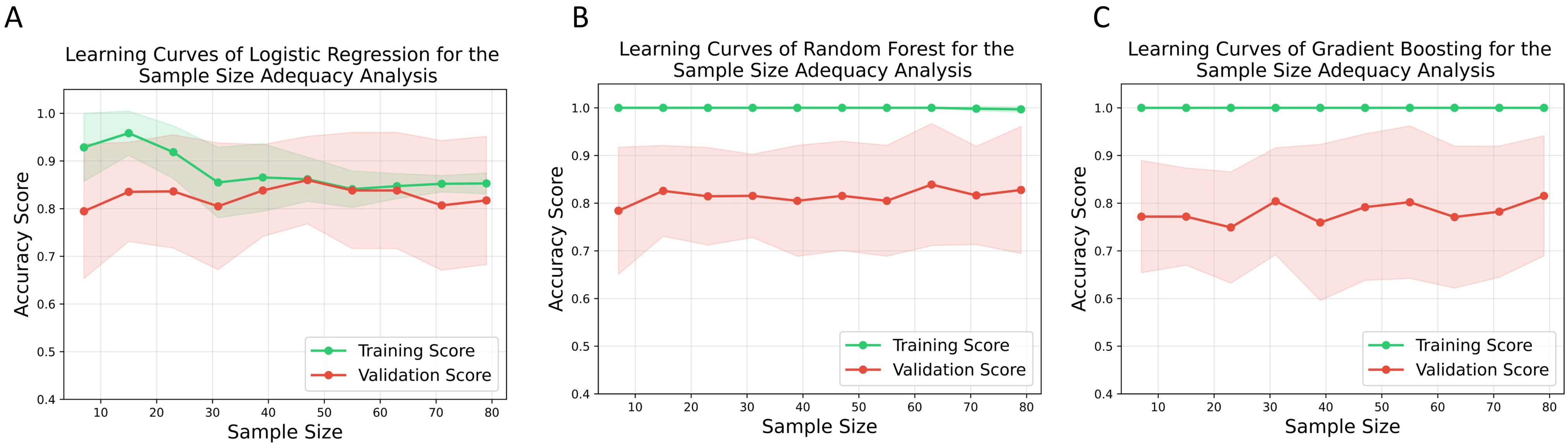


**S4 Figure**: **Learning curve analysis for different ML classifiers for the NCa vs. Ca predictor. A.** Learning curves for the logistic regression (LR) model. High validation scores and Low variance between the training and validation scores indicate that LR can be an adequate classifier. **B.** Learning curves for the random forest (RF) method. High variance between the training and validation scores shows that RF is not an adequate classifier. **C.** Learning curves for the gradient boosting (GB) method. High variance between the training and validation scores shows that GB is not an adequate classifier. In each analysis, sample proportions ranging from 10% to 100% of the training dataset were sampled multiple times. A stratified k-fold cross-validation was used to obtain the training (shown in green) and validation (shown in red) performance curves across varying training dataset sample sizes. Shaded regions represent standard deviation across 8-fold cross-validation.

**S2 Table:** The number of patients with missing data in the Ca vs. PCa cohort with 156-patients and 37 biomarkers

| | Missing |
|---|---|
| ENA.78 | 0 |
| IFN.y | 0 |
| IL.10 | 1 |
| IL.6 | 0 |
| IL.8 | 0 |
| MCP.1 | 0 |
| MDC | 0 |
| MIP.1a | 5 |
| TNF.a | 0 |
| C.peptide | 1 |
| G.CSF | 0 |
| IL.22 | 7 |
| Insulin | 3 |
| Leptin | 3 |
| MIP.3a | 10 |
| GRO.a | 0 |
| HGF | 0 |
| MMP.2 | 2 |
| Adiponectin | 0 |
| CRP | 0 |
| GDF.15 | 0 |
| TIMP.1 | 0 |
| TGF.B2 | 18 |
| TGF.B1 | 0 |
| PPAR.y | 1 |
| HIF.1a | 11 |
| Laminin | 0 |
| HbA1c | 0 |
| CA19.9 | 14 |
| Glucose | 5 |
| HDL | 0 |
| CCK | 1 |
| LDL | 0 |
| Triglyceride | 18 |
| Albumin | 2 |
| Lumican | 0 |
| ZAG | 0 |

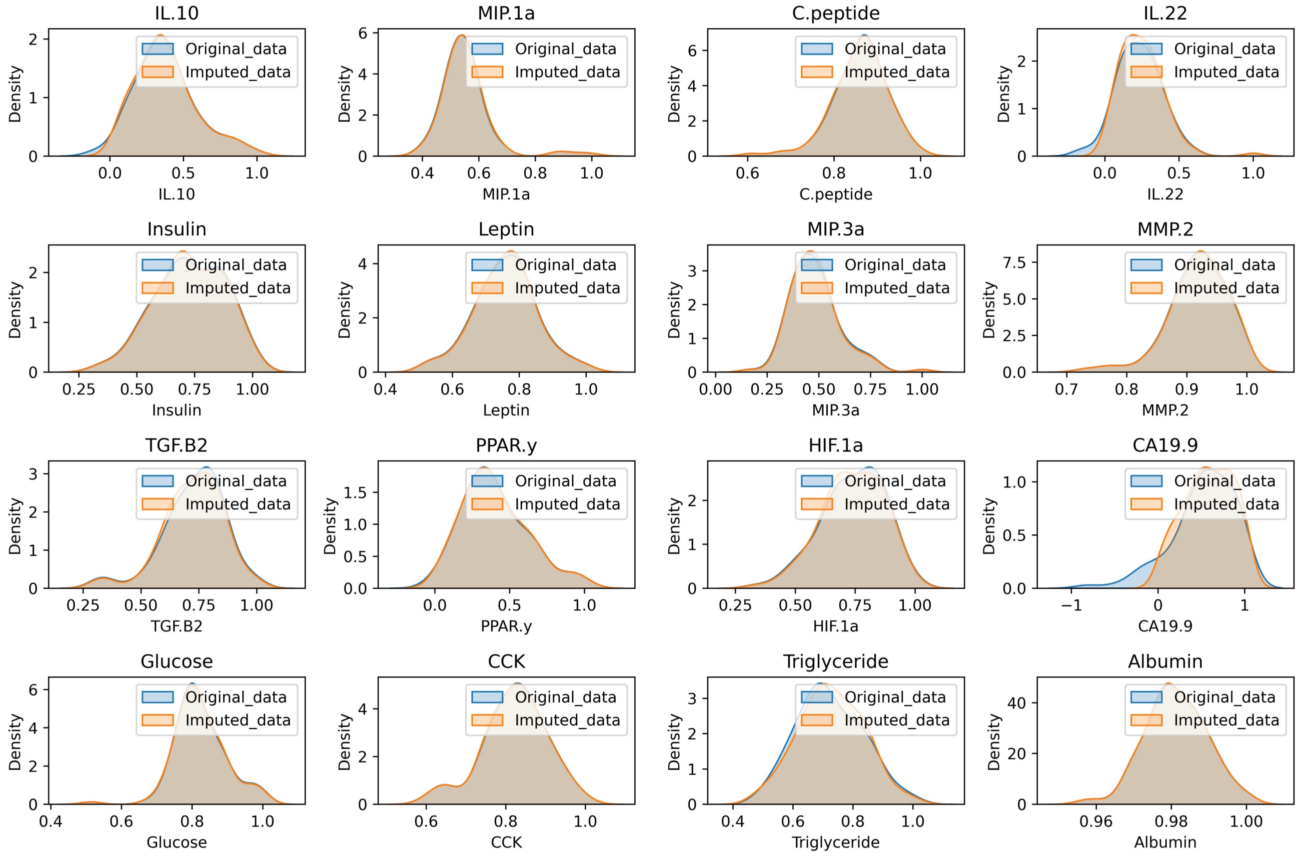


**S5 Figure:** Density plots of the training dataset (blue) and the imputed training dataset (orange) in 16 biomarkers with missing data for the Ca vs. PCa predictor.

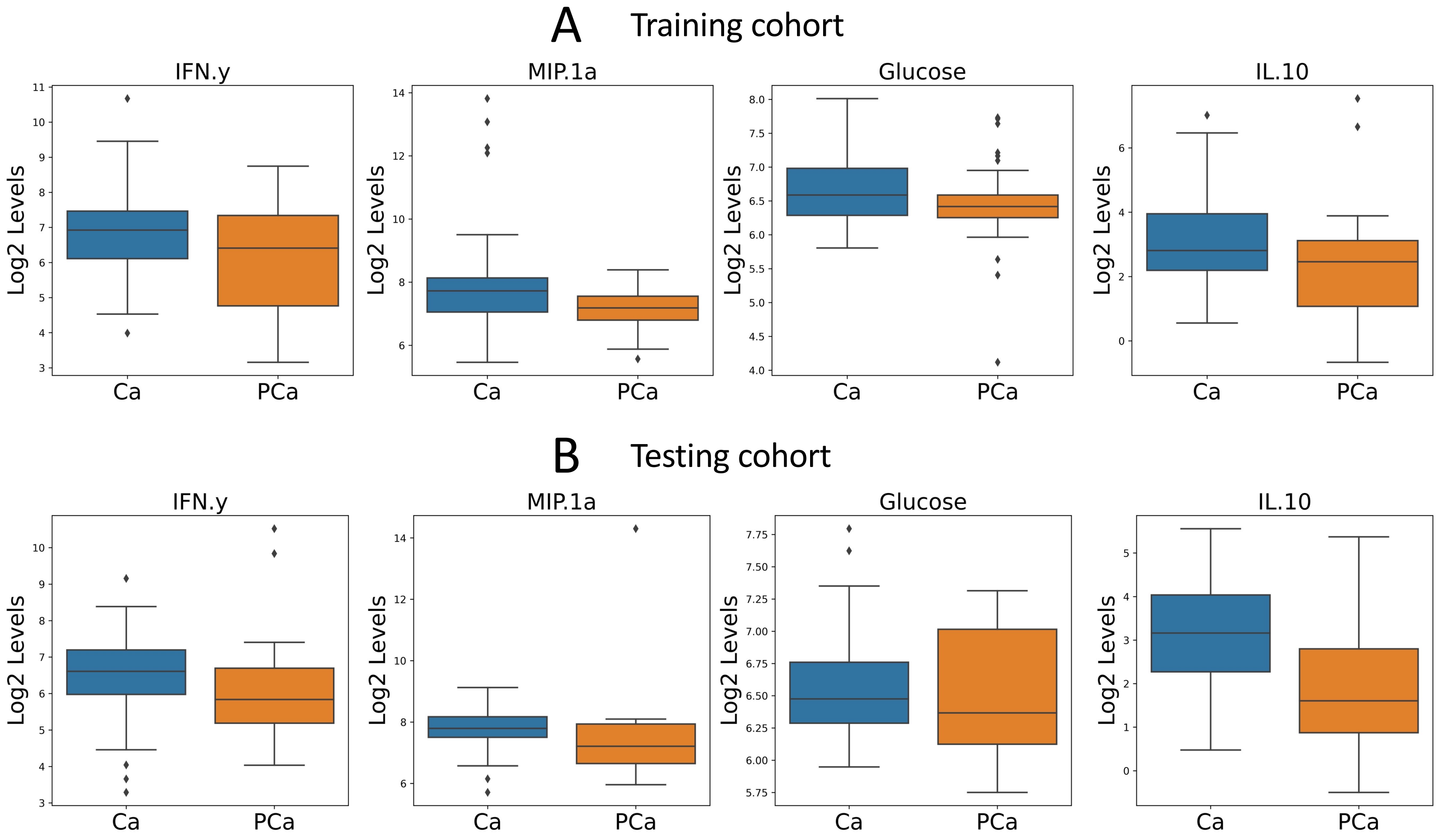


**S6 Figure: Distributions of robust predictive biomarkers for the Ca vs. PCa predictor.** Distributions of the Ca (blue) and PCa (orange) classes for the training **A** and testing **B** cohorts for the four robust predictive biomarkers.

A B C

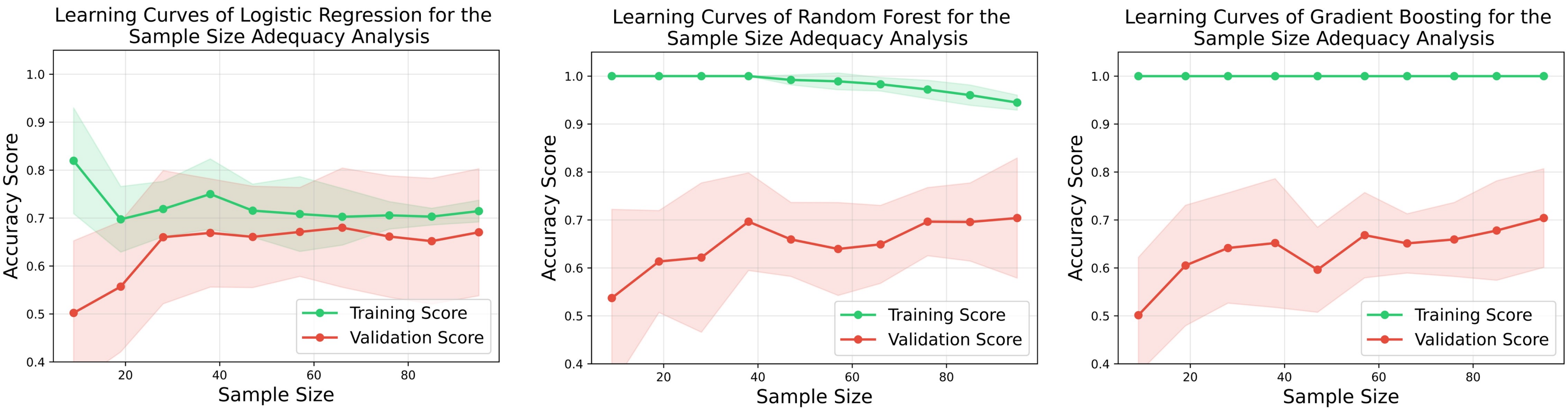


**S7 Figure**: **Learning curve analysis for different ML classifiers for the Ca vs. PCa predictor. A.** Learning curves for the logistic regression (LR) model. Low variance between the training and validation scores indicate that LR can be an adequate classifier. **B.** Learning curves for the random forest (RF) method. High variance between the training and validation scores shows that RF is not an adequate classifier. **C.** Learning curves for the gradient boosting (GB) method. High variance between the training and validation scores shows that GB is not an adequate classifier. In each analysis, sample proportions ranging from 10% to 100% of the training dataset were sampled multiple times. A stratified k-fold cross-validation was used to obtain the training (shown in green) and validation (shown in red) performance curves across varying training dataset sample sizes. Shaded regions represent standard deviation across 8-fold cross-validation.

**S3 Table:** The number of patients with missing data in the NCa vs. PCa cohort with 81-patients and 37 biomarkers

| | Missing |
|---|---|
| ENA.78 | 0 |
| IFN.y | 0 |
| IL.10 | 1 |
| IL.6 | 0 |
| IL.8 | 0 |
| MCP.1 | 0 |
| MDC | 0 |
| MIP.1a | 3 |
| TNF.a | 0 |
| C.peptide | 1 |
| G.CSF | 0 |
| IL.22 | 6 |
| Insulin | 2 |
| Leptin | 0 |
| MIP.3a | 10 |
| GRO.a | 0 |
| HGF | 0 |
| MMP.2 | 0 |
| Adiponectin | 0 |
| CRP | 0 |
| GDF.15 | 0 |
| TIMP.1 | 0 |
| TGF.B2 | 8 |
| TGF.B1 | 0 |
| PPAR.y | 0 |
| HIF.1a | 3 |
| Laminin | 0 |
| HbA1c | 0 |
| CA19.9 | 5 |
| Glucose | 1 |
| HDL | 0 |
| CCK | 0 |
| LDL | 0 |
| Triglyceride | 11 |
| Albumin | 1 |
| Lumican | 0 |
| ZAG | 0 |

#### S8 Figure

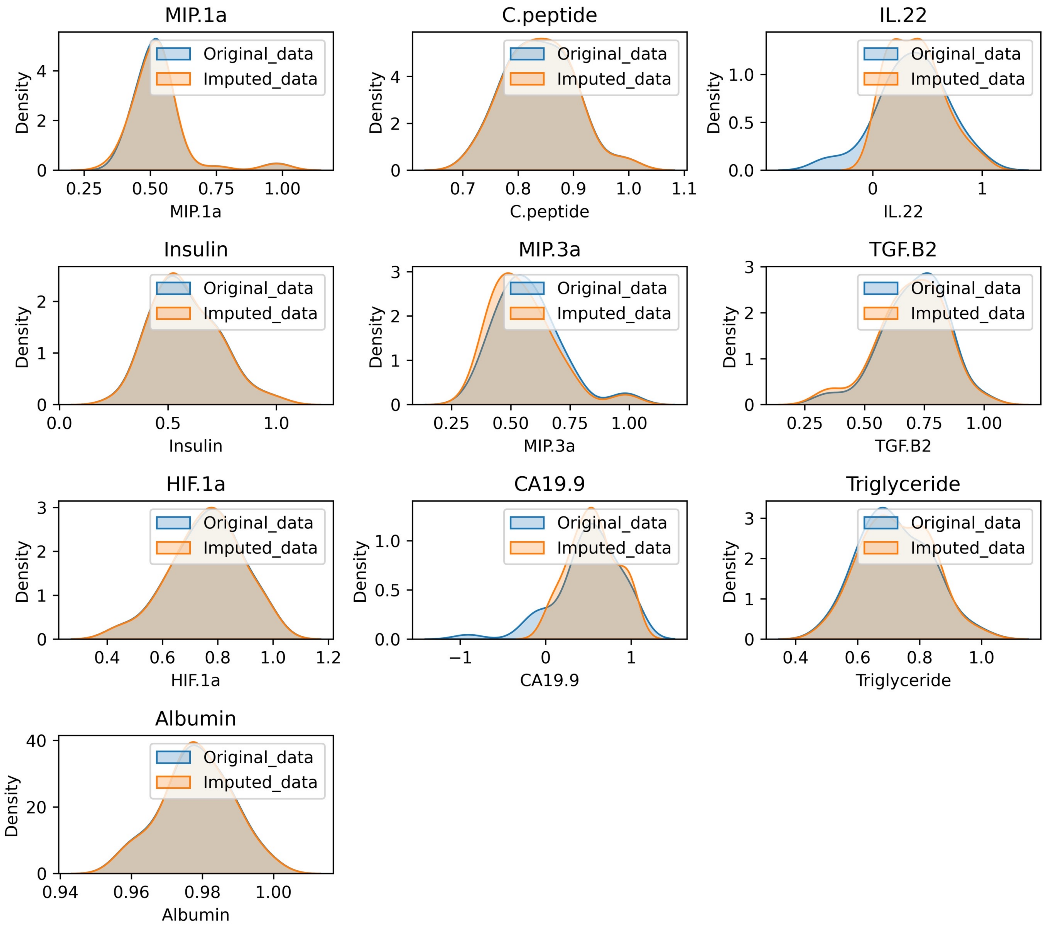


**S8 Figure:** Density plots of the training dataset (blue) and the imputed training dataset (orange) in 10 biomarkers with missing data for the NCa vs. PCa predictor.

#### S9 Figure

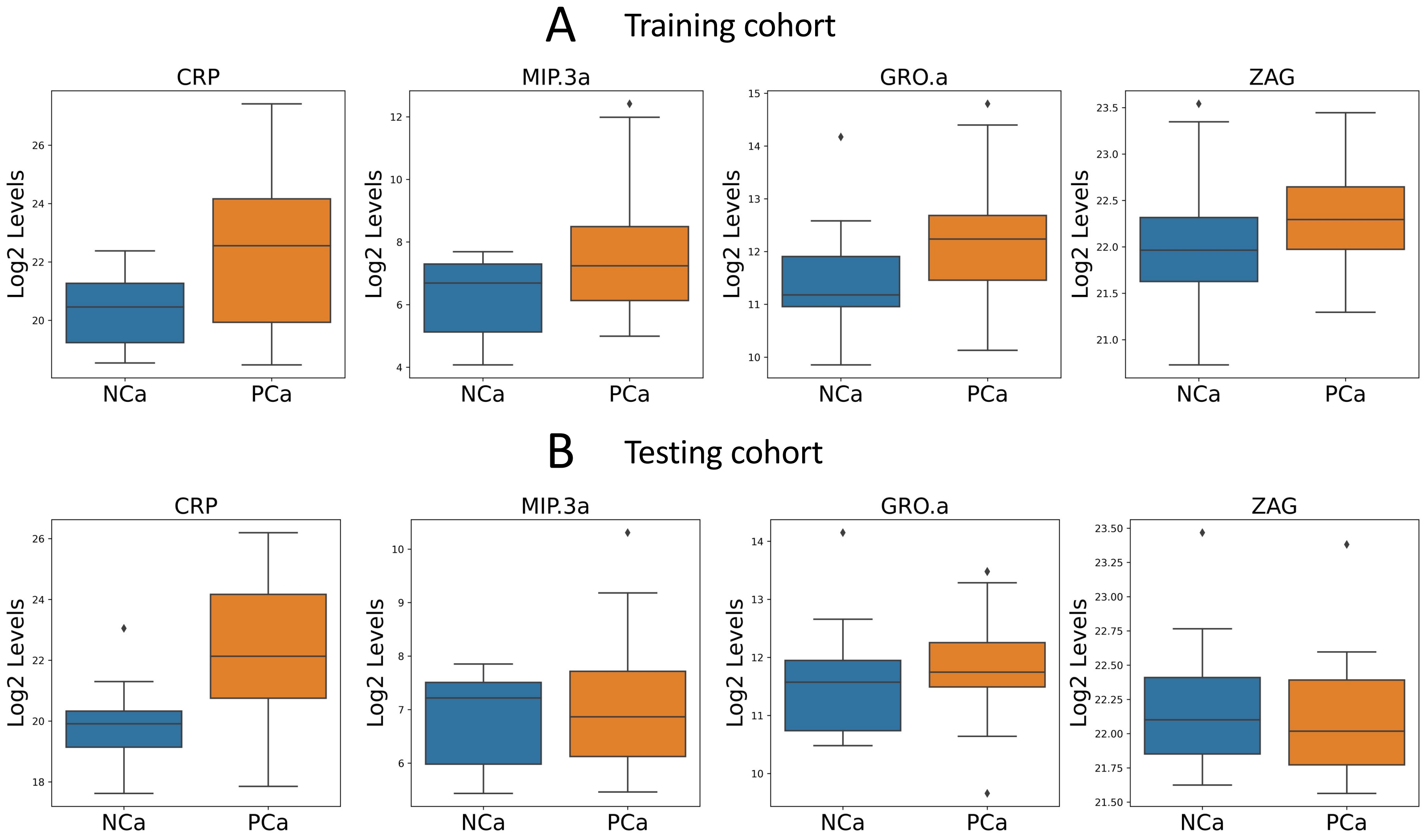


**S9 Figure: Distributions of robust predictive biomarkers for the NCa vs. PCa predictor.** Distributions of the NCa (blue) and PCa (orange) classes for the training **A** and testing **B** cohorts for the four robust predictive biomarkers.

### S10 Figure

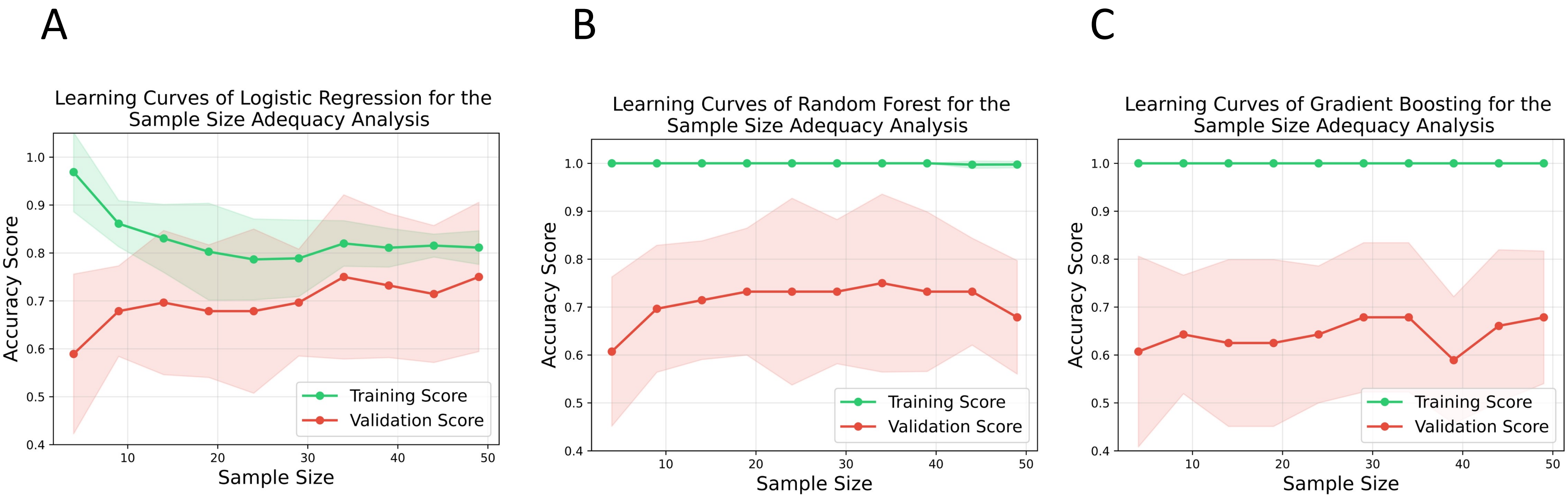


**S10 Figure**: **Learning curve analysis for different ML classifiers for the NCa vs. PCa predictor. A.** Learning curves for the logistic regression (LR) model. Low variance between the training and validation scores indicate that LR can be an adequate classifier. **B.** Learning curves for the random forest (RF) method. High variance between the training and validation scores shows that RF is not an adequate classifier. **C.** Learning curves for the gradient boosting (GB) method. High variance between the training and validation scores shows that GB is not an adequate classifier. In each analysis, sample proportions ranging from 10% to 100% of the training dataset were sampled multiple times. A stratified k-fold cross-validation was used to obtain the training (shown in green) and validation (shown in red) performance curves across varying training dataset sample sizes. Shaded regions represent standard deviation across 8-fold cross-validation.

### S1 Algorithm

**Algorithm 1:** Determine Optimal Threshold using RBF-SVM and MCC

**Input:** Labeled training data $\mathcal{D}_{\text{train}} = (\mathbf{X}, \mathbf{Y})$, where $\mathbf{X} = [\mathbf{X}_1, \mathbf{X}_2, \cdots, \mathbf{X}_M]^\intercal$, each $\mathbf{X}_i = \left(\mathbf{x}_i^1, \mathbf{x}_i^2, \cdots, \mathbf{x}_i^P\right)$, and $\mathbf{Y} = [y_1, y_2, \cdots, y_M]^\intercal$, where each $y_i \in \{-1, 1\}$

. **Output:** Decision threshold $\theta^*$ that maximizes classification performance

Train a radial basis function (RBF) kernel SVM classifier $\mathcal{M}$ on $\mathcal{D}_{\text{train}}$ with optimal hyperparameters

For each input $\mathbf{X}_i$, compute the predicted class probability $\hat{p}_i = \mathbb{P}_{\mathcal{M}}(y_i = 1 \mid \mathbf{X}_i)$

**for** *threshold* $\theta$ *from* 0.01 *to* 0.99 *in steps of* 0.001 **do**

Convert probabilities to binary predictions using:

$$\hat{y}_i^{(\theta)} = \begin{cases} 1 & \text{if } \hat{p}_i \geq \theta \quad \text{(`Postive Class')} \\ -1 & \text{otherwise} \quad \text{(`Negative Class')} \end{cases}$$

Compute the Matthews correlation coefficient (MCC) between the predicted labels $\hat{\mathbf{Y}}^{(\theta)} = \left[\hat{y}_1^{(\theta)}, \hat{y}_2^{(\theta)}, \cdots, \hat{y}_M^{(\theta)}\right]^\intercal$ and the true labels $\mathbf{Y}$

Identify the threshold that yields the best performance:

$\theta^* \leftarrow \arg\max_\theta \text{MCC}(\hat{\mathbf{Y}}^{(\theta)}, \mathbf{Y})$

Apply $\theta^*$ to classify instances in the test set using probability outputs from $\mathcal{M}$

**S1 Algorithm:** Algorithm for determining the optimal decision threshold for radial basis-kernel function for the support vector machine (RBF-SVM) and the Matthews correlation coefficient (MCC) methods.
